# Antibody neutralization to SARS-CoV-2 and variants after one year in Wuhan

**DOI:** 10.1101/2021.06.16.21258673

**Authors:** Qianyun Liu, Qing Xiong, Fanghua Mei, Chengbao Ma, Zhen Zhang, Bing Hu, Junqiang Xu, Yongzhong Jiang, Faxian Zhan, Xianying Chen, Ming Guo, Xin Wang, Yaohui Fang, Shu Shen, Yingle Liu, Fang Liu, Li Zhou, Ke Xu, Changwen Ke, Fei Deng, Kun Cai, Huan Yan, Yu Chen, Ke Lan

**Affiliations:** State Key Laboratory of Virology, Institute for Vaccine Research and Modern Virology Research Center, College of Life Sciences, Wuhan University, Wuhan, China; Hubei Provincial Center for Disease Control and Prevention, Wuhan, China; National Virus Resource Center, Wuhan Institute of Virology, Chinese Academy of Sciences; Guangdong Provincial Center for Disease Control and Prevention, Guangzhou, China

## Abstract

Most COVID-19 patients can build effective humoral immunity against SARS-CoV-2 after recovery(*1, 2*). However, it remains unknown how long the protection can maintain and how efficiently it can protect people from the reinfection of the emerging SARS-CoV-2 variants. Here we evaluated the sera from 248 COVID-19 convalescents around one year post-infection in Wuhan, the earliest epicenter of SARS-CoV-2. We demonstrated that the SARS-CoV-2 immunoglobulin G (IgG) maintains at a high level and potently neutralizes the infection of the original strain (WT) and the B.1.1.7 variant in most patients. However, they showed varying degrees of efficacy reduction against the other variants of concern (P.1, B.1.525, and especially B.1.351) in a patient-specific manner. Mutations in RBD including K417N, E484K, and E484Q/L452R (B.1.617) remarkably impair the neutralizing activity of the convalescents’ sera. Encouragingly, we found that a small fraction of patients’ sera showed broad neutralization potency to multiple variants and mutants, suggesting the existence of broadly neutralizing antibodies recognizing the epitopes beyond the mutation sites. Our results suggest that the SARS-CoV-2 vaccination effectiveness relies more on the timely re-administration of the epitope-updated vaccine than the durability of the neutralizing antibodies.

## Main

The SARS-CoV-2 emerged more than a year ago rapidly swept across the world and developed into a long-lasting COVID-19 pandemic with devastating impacts(*3, 4*). The situation leads to the perpetual mutation of SARS-CoV-2 with numerous variants emerging around the world. In addition to the initial D614G mutation, the viral spike protein undergone antigenic drift and produced several variants of concern associated with local outbreaks, such as B.1.1.7 (United Kingdom)(*5*), B.1.351 (South African)(*6*), P.1 (Brazil, also known as B.1.1.28.1)(*7*), B.1.525 (Nigeria)(*8*) and B.1.617 (India)(*9*).

So far, due to the lack of sufficient evidence about how long the anti-SARS-CoV-2 protective immunity induced by prior infection or vaccination can maintain, the optimal interval between the vaccinations remains to be determined(*10, 11*). Additionally, increasing evidence showing these variants of concern, especially the South African strain B.1.351(*12, 13*), remarkably increased resistance to the neutralizing antibodies, which largely due to the mutations in the RBD region interacting with human angiotensin-converting enzyme 2 (hACE2)(*3*). It is imperative to figure out the durability and cross-variant protection of SARS-CoV-2 neutralizing antibodies elicited by previous infection or vaccination based on the original strain.

Wuhan is the first known epicenter of the SARS-CoV-2 outbreak. Almost all patients in this city were infected by the original strain before the city reopen and the epidemic here had been well controlled thereafter before the emergence of variants(*14*). In this study, sera from convalescents infected around one year ago in Wuhan were collected to investigate the SARS-CoV-2 antibody durability and the cross-variant protection over time. Results from Enzyme-linked immunosorbent assay (ELISA) and pseudovirus neutralization assay demonstrated that the SARS-CoV-2 RBD-specific IgG antibodies remain detectable in most patients, which potently inhibit the infection of the SARS-CoV-2 pseudovirus (WT-D614G and B.1.1.7), but to a different extent compromised to other tested variants (P.1, B.1.525 and especially B.1.351) and mutations (K417N, L452R, E484K, and E484Q/L452R). The decline of neutralizing activity was confirmed by authentic viral neutralization assays (WT, B.1.1.7 and B.1.351). Of note, we showed eight individuals developed relatively broad immunity against the tested variants, supporting the presence of broadly neutralizing epitopes. Overall, these results provided important implications for the control of the pandemic caused by the constantly emerging SARS-CoV-2 variants.

### Durability and neutralizing activity

To evaluate the durability and neutralizing activity of SARS-CoV-2 antibodies after infection, we collected sera and clinical records from 248 COVID-19 patients infected around one year ago (11-12 months after symptom onset) in Wuhan. Firstly, a rapid ELISA (colloidal gold) test for SARS-CoV-2-S IgM/IgG antibody was applied. 79 % (197/248) of the sera tested positive for anti-SARS-CoV-2 IgG, and 2.0 % (5/248) tested positive for anti-SARS-CoV-2 IgM (Table S1). Conventional ELISA assays were then conducted to determine the anti-RBD IgG level and the competitive ability of ACE2 to RBD-binding of the sera. As we set OD450=0.31 (the mean OD450 value + 3 SD of uninfected people’s sera) as the cut-off value, 91 % (225/248) tested positive for anti-SARS-CoV-2-RBD IgG (**Fig. 1A**). Our results also showed that the competitive ability is highly consistent with the anti-RBD IgG level (**Fig. 1A-E**, Table S1), indicating most RBD-targeting antibodies in the patients can interfere with the interaction between ACE2 and RBD. However, the anti-RBD IgG level and the neutralizing activity of the sera showed no statistically significant difference in patients with different severity (Fig.1a-b), gender (Fig.1c-d), and age (Fig. 1e).

**Fig. 1:**
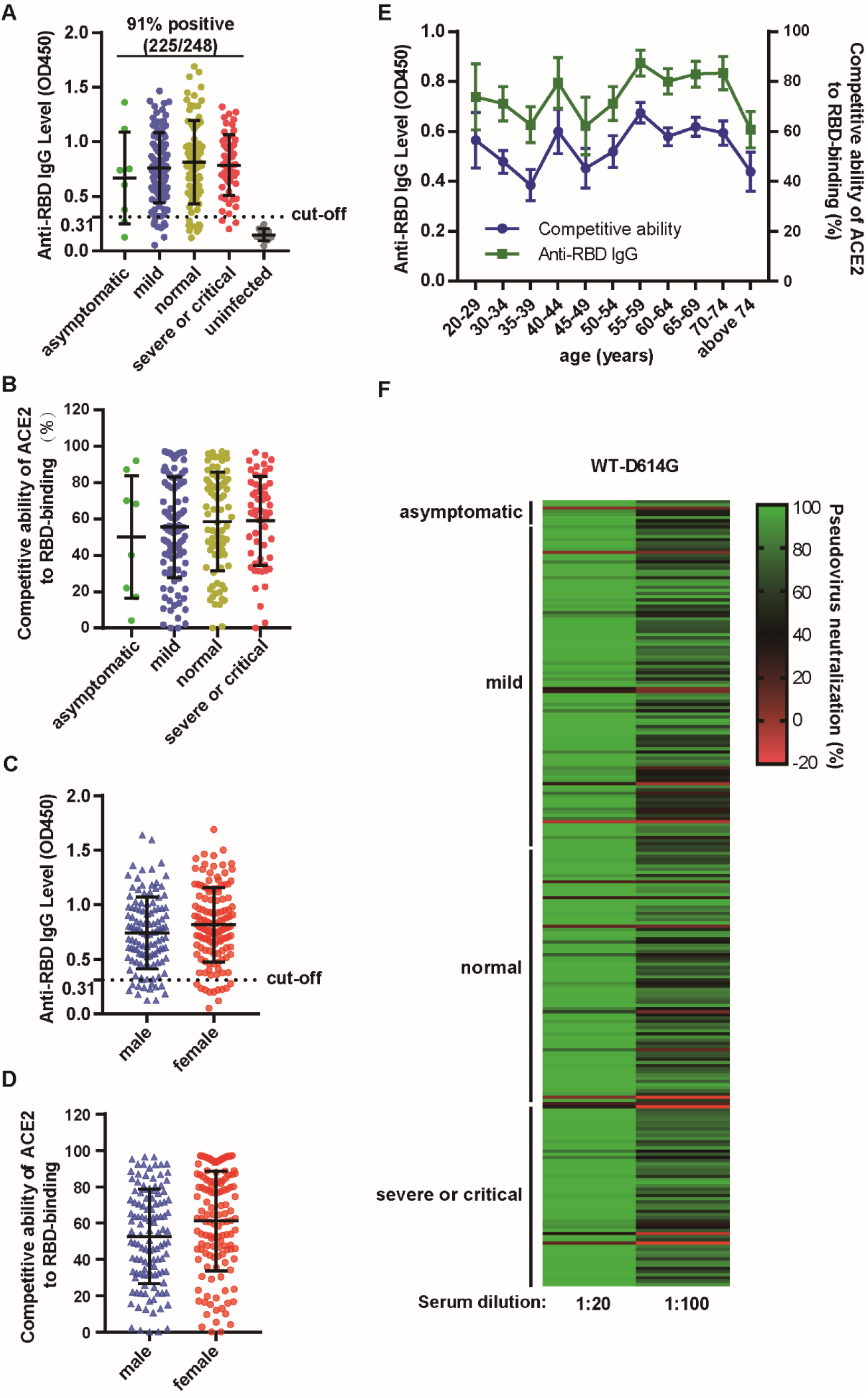
The anti-RBD IgG level and neutralizing activity of sera from COVID-19 patients in Wuhan one-year after recovery. **a-e**, Results of ELISA measuring the 248 convalescent COVID-19 patients’ sera reactivity to SARS-CoV-2-RBD. The anti-RBD IgG level of the indicated groups **(a, c)**. The competitive ability of ACE2 to RBD-binding of the indicated groups **(b, d). Comparison of t**he anti-RBD IgG and the competitive ability of ACE2 to RBD-binding in patients with different age (**e**). **f**, The neutralizing activity (WT-D614G) of the sera from convalescents with different severity was tested with the indicated dilution folds. Mean ± s.d. are shown in **a-d**. Mean ± s.e.m are shown in **e**. Statistical significance was determined using two-tailed Mann–Whitney U-tests, and no significant difference was found (*P* > 0.05). The horizontal dotted lines on **a** and **c** indicate the cut-off value (0.45, three times of t the mean OD450 value of uninfected people’s sera).

The neutralizing activities of the sera were tested with SARS-CoV-2 pseudovirus (WT-D614G) produced based on a replication-deficient VSV pseudotyping system (VSV-dG-Luc) and a BHK-21 cell line stably expressing hACE2 (BHK-21-hACE2). 234 sera (94 %) showed more than 50 % neutralization when diluted at 20-fold (NT_50_ > 20), and 195 samples (79 %) showed 50 % neutralization under 100-fold dilution (NT_50_ > 100). No statistically significant difference was observed between pseudovirus neutralizing activity and the disease severity (Fig. 1f). These results indicate that most patients could build and maintain an effective humoral immunity against SARS-CoV-2 for at least one year.

### Cross-variant protection

Recently, SARS-CoV-2 variants with multiple mutations in S protein have emerged in the United Kingdom (B.1.1.7), Brazil (P.1), Nigeria (B.1.525), South Africa (B.1.351), United states (B.1.427/B.1.429)(*15*), India (B.1.617), *etc*. We first tested the neutralizing activity of the top 180 potent sera in Fig. 1f by the pseudoviruses of four variants, with the WT-D614G as a control (Fig. 2a). When tested at 100-fold dilution, the mean neutralization efficiency of the sera against WT-D614G is 65 %. However, the efficiency reduced to 59 % for B.1.1.7, 48 % for P.1, 43 % for B.1.525, and 35 % for B.1.351 (Fig. 2b). Interestingly, we found that a small fraction of the patients showed cross-variant neutralizing activity. Accordingly, we selected eight representative sera from those patients (group 1, relatively resistant) for further verification by NT_50_ determination, with another eight sera (group 2, relatively sensitive) for comparison. As expected, the sera in group 1 showed only a slight decrease in NT_50_ against the tested variants compared with WT-D614G (1.26∼2.57 folds), while the sera in group 2 showed much higher sensitivity (1.83∼8.10 folds). The NT_50_ data further confirmed that the B.1.351 is the top immune escape variant among the tested strains, followed by B.1.525, P.1, and B.1.1.7, respectively (Fig. 2c-d, and Extended data 1a).

**Fig. 2:**
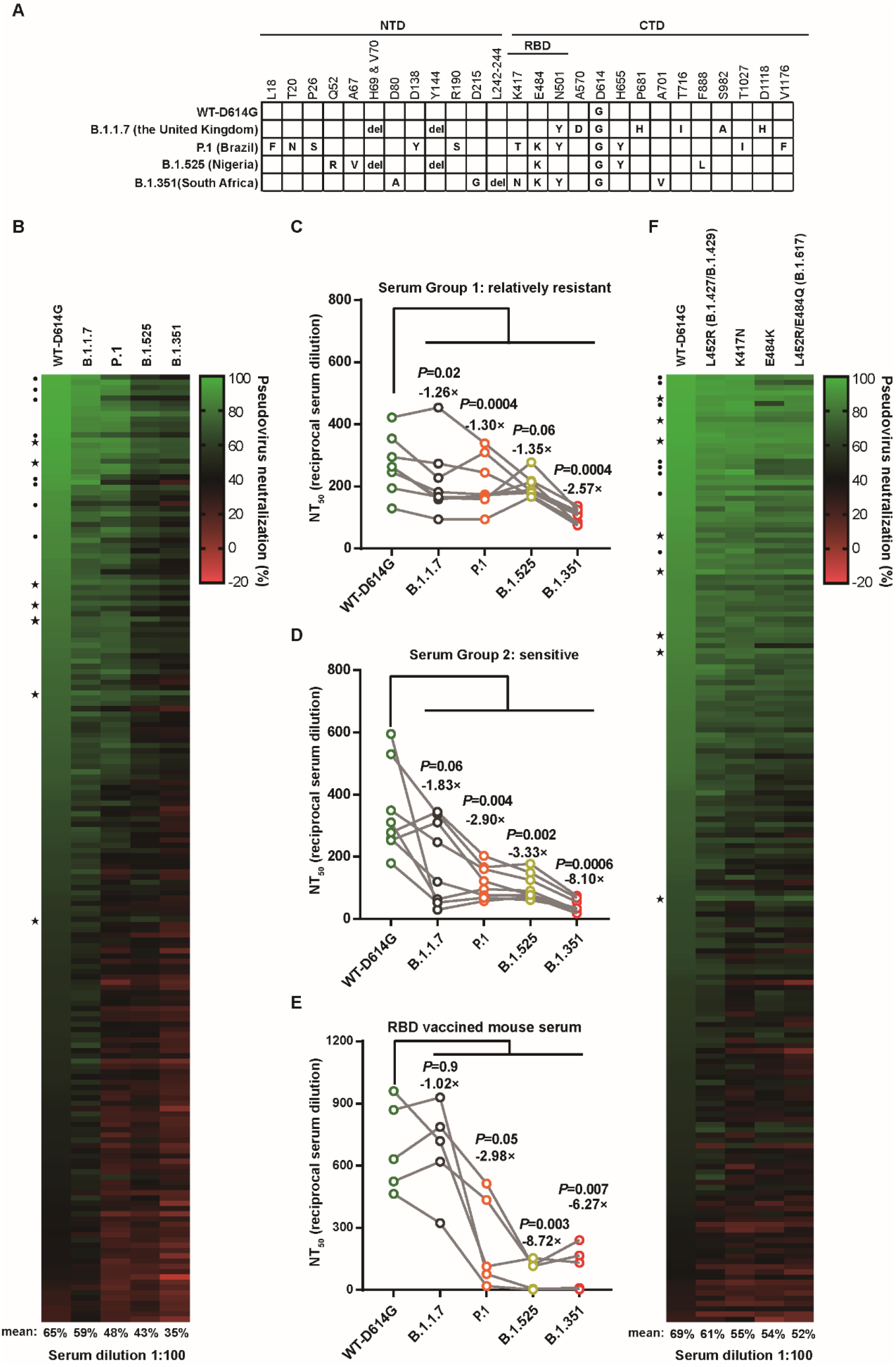
The neutralizing activity of the convalescents’ sera to SARS-CoV-2 variants. **a**, The amino acid alterations in the S protein of SARS-CoV-2 variants tested in this study. **b**, The neutralizing activity of convalescents’ sera at 100-fold dilution against pseudoviruses bearing S proteins from the indicated variants with WT-D614G as a control. **c-e**, Paired analysis of NT_50_ values of convalescents’ sera or RBD vaccinated mice sera against WT-D614G and variants. Serum group 1: relatively resistant **(c)**. Serum group 2: sensitive **(d)**. RBD-vaccined mice sera **(e)**. The serum group 1 and 2 were selected according to their tolerance to mutations in B.1.351. Statistical significance was determined using paired two-tailed t-tests. *P* values and mean fold changes in NT_50_ values compared with WT-D614G are indicated. **f**, The neutralizing activity of convalescents’ sera at 100-fold dilution against pseudoviruses bearing different S with indicated mutations. •, the relatively resistant serum sample.⋆, the sensitive serum sample.

We further tested whether the immune escape of the tested variants is mainly attributed to the reduced neutralizing activity of SARS-CoV-2 RBD-specific antibodies. Five C57BL/6 mice were immunized three times with WT SARS-CoV-2 RBD human IgG-Fc recombinant protein. Seven weeks later, the mice sera were collected and tested by pseudovirus neutralization assay. Similar to most of the convalescents’ sera, the sera from RBD protein immunized mice were sensitive to B.1.525, P.1, and B.1.351, but not B.1.1.7, which is reasonable as B.1.1.7 only have one mutation (N501Y) in the RBD region, a mutation without prominent immune escape effect(*12, 16*) (Fig. 2e). These results showed that the immune escape ability of the variants can be largely attributed to the mutations in the RBD region.

To define the contribution of the critical mutations in RBD region to immune escape, we produced pseudovirus bearing SARS-CoV-2 S protein (D614G) with K417N, L452R, E484K, and L452R/E484Q mutations, respectively. Notably, L452R and L452R/E484Q are two critical mutations in the B.1.427/B.1.429 (The United States) and B.1.167 (India) variant, respectively. We tested the neutralizing activity of the 180 sera by these mutants with the WT-D614G as a control. As shown in Fig. 2f, when tested at 100-fold dilution, the mean neutralization efficiency of the sera against WT-D614G is 69 %. However, the efficiency reduced to 61 % for L452R, 55 % for K417N, 54 % for E484K, and 52 % for L452R/E484Q. This result indicates that the RBD mutations are responsible for the reduction of cross-variant neutralization among the variants of concern, especially mutations on E484 and K417.

### Verification by authentic viruses

To confirm whether the neutralization results are consistent between the SARS-CoV-2 pseudovirus and the authentic virus, we first conducted the authentic SARS-CoV-2 virus (WT) neutralization assay to verify the 30 representative sera showing high, medium, and low neutralizing activities in Fig. 1f, respectively. The SARS-CoV-2 nucleocapsid (N) protein immunofluorescence assay showed consistent results with the pseudovirus neutralization assay (Fig. 3a).

**Fig. 3:**
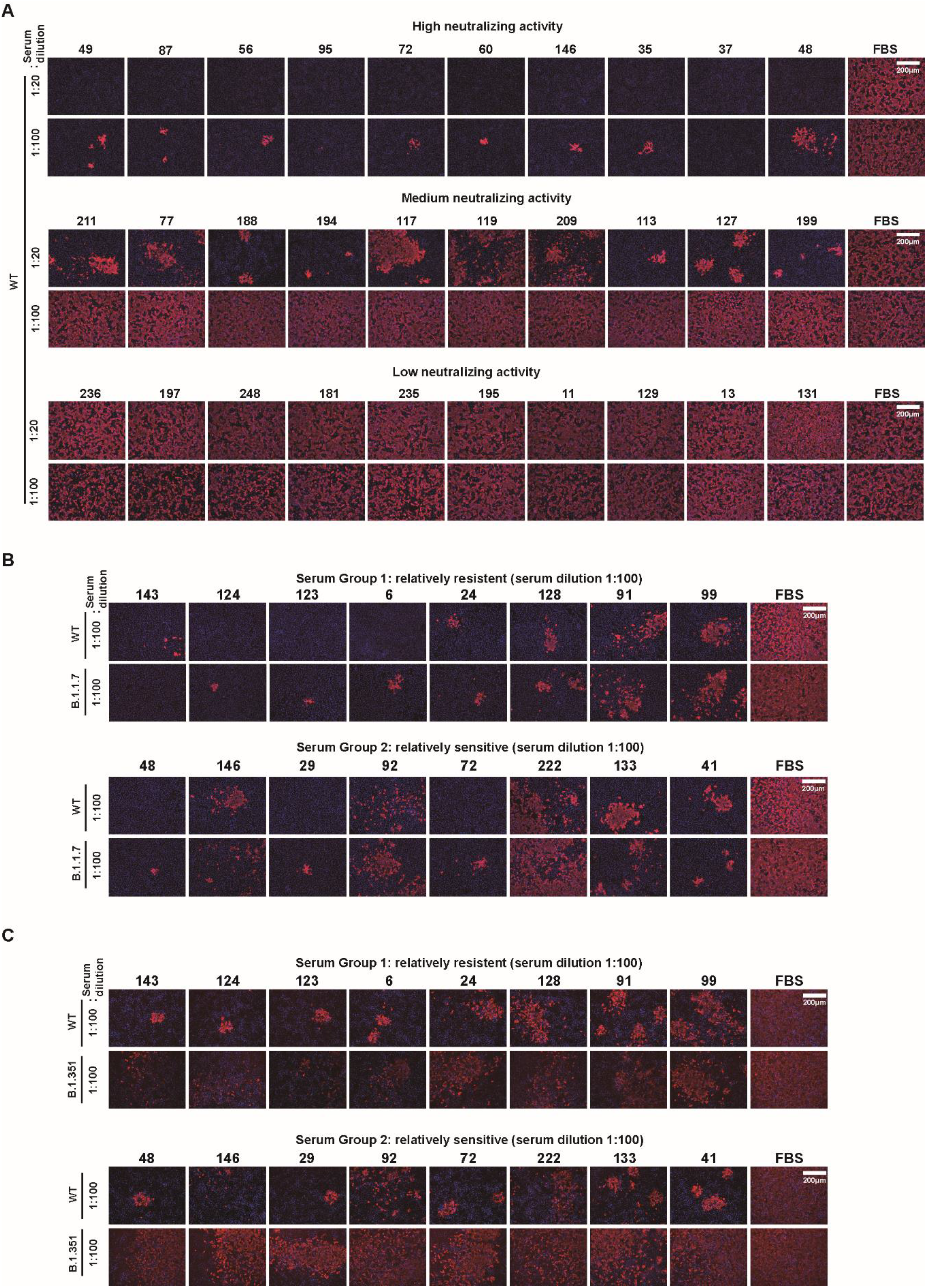
The neutralizing activity of the convalescents’ sera to authentic SARS-CoV-2 viruses. **a**, The neutralizing activity of 30 representative sera against authentic SARS-CoV-2 WT strain at 20-fold and 100-fold dilution. The sera were grouped based on their neutralizing activity to WT-D614G pseudoviruses. **b-c**, The neutralizing activity of the selected sera (relatively resistant or sensitive against B.1.351 pseudovirus) against the indicated authentic SARS-CoV-2 viruses. SARS-CoV-2 N protein (red) in the infected cells was detected through immunofluorescence assay at 24 h post-infection. The nucleus was stained blue. Scale bar, 200 μm. The serum sample number were indicated above the pictures.

We further verified the neutralizing activity of two groups of patients in Fig. 2 c-d (relatively sensitive and relatively resistant) by three authentic SARS-CoV-2 strains, including the WT strain and the B.1.1.7 and B.1.351 variant strains. The sera were diluted 100-fold, pre-incubated with these viruses, and then applied to Vero E6 cells for infection neutralization assay. As shown in Fig. 3b, the neutralizing activity of the sera against the B.1.1.7 strain is similar to the WT strain but with a slight decrease. As for the B.1.351 strain, the group 1 sera (relatively resistant) efficiently neutralized the infection of both the WT and the B.1.351 strains. In contrast, the group 2 sera (relatively sensitive) showed a remarkable reduction of neutralizing activity against the B.1.351 strain compared with the WT strain (Fig. 3c). Again, these results are also consistent with the results from pseudovirus neutralization assays.

## Discussion

Previous studies show the protective immunity against seasonal coronaviruses (HCoV-NL63, HCoV-229E, HCoV-OC43 and HCoV-HKU1) may last only 6 to 12 months(*17*), and the titer of SARS-CoV-2 antibody has been reported to decline rapidly in the first several months(*10*), raising the concern about the durability of SARS-CoV-2 neutralizing antibodies elicited by either previous infection or vaccination. Additionally, SARS-CoV-2 variants are constantly emerging during the COVID-19 pandemic, greatly challenging the development and application of effective neutralizing antibodies and vaccines. It is imperative to investigate the durability and cross-variant protection of the anti-SARS-CoV-2 humoral immunity. To this end, we collected convalescent sera in Wuhan in consideration of the several unique advantages over the samples collected elsewhere: (1) Wuhan is the earliest reported epicenter of COVID-19 with most patients infected more than one year ago; (2) The patients were all infected by the original WT strain without the interference of potential multi-variants infection; (3) So far, most administrated vaccines globally were designed based on the sequences of the original strain from Wuhan.

Firstly, we report that the effective neutralizing antibodies against SARS-CoV-2 could last at least one year in most COVID-19 convalescents, while no statistical significance was observed among different severity, gender, or age (Fig. 1a-e). These results indicate that most people including asymptomatic and elderly patients could build and maintain effective anti-SARS-CoV-2 humoral immunity, with a few exceptions in each group for unknown reasons (Fig. 1). We speculate this phenotype might be attributed to the individual immune difference that deserves further study.

Another particular concern is the cross-variant protective efficacy of the anti-SARS-CoV-2 humoral immunity. Our study provides additional evidence that most sera collected from convalescents in Wuhan, even for one year post-infection, can still efficiently neutralize the infection of B.1.1.7 variant. However, a significant decline of efficacy was observed on P.1, B.1.525, and B.1.351 (Fig. 2b). B.1.351 displayed the most prominent immune escape ability among these tested variants, consisting with the previous reports(*12, 13, 18*). Encouragingly, sera from a few individuals showed resistance to all tested variants (Fig. 2c), indicating the presence of effective and broadly neutralizing antibodies. It brings hope for the identification of ideal broad neutralization epitopes essential for the development of antibody therapy and next-generation vaccines.

We next sought to characterize the critical restudies that contribute to the immune escape. Based on previous studies and our neutralization results from RBD vaccinated mouse sera, we focus on the four mutation sites in RBD of the tested variants in this study (Fig. 2a). N501Y is the only mutation in B.1.1.7 RBD, but this variant did not show remarkable immune escape in both our (Fig. 2e) and prior studies(*19, 20*). Therefore, we tested several mutations in the other three critical sites, including K417N, L452R, E484K, and L452R/E484Q. The neutralization assay demonstrated that the L452R/E484Q (two key mutations of B.1.617) showed the strongest immune escape, followed by E484K, K417N, and L452R. The mean sera neutralization efficiency (Fig. 2b and 2f) demonstrated that the critical RBD mutations play a major role in the immune escape of the variants. Importantly, the cross-variant protective sera from the group 1 convalescents can also tolerate these critical mutations (Fig. 2f).

We do acknowledge some limitations of our studies. First, we did not determine the NT_50_ of all the sera because of the limitation of the sample volume, especially for those with lower neutralizing activity. Second. we did not include all the possible mutations that can appear in the B.1.617 and its sub-lineages in consideration of the sequence complexity of the variants circulating in India. Thus, future studies could be done to investigate whether other mutations on spike proteins can result in a leap of the immune escape ability of B.1.617 and other variants.

To our knowledge, the current study provides the first direct evidence that the potent anti-SARS-CoV-2 humoral immunity can last for at least one year in most convalescents to protect them against the original virus (WT or WT-D614G), which means the annually re-administration might be a feasible vaccination strategy to maintain the anti-SARS-CoV-2 humoral immunity. However, the convalescents and the vaccinated people are gradually losing protection against the constantly emerging immune escape variants. It remains to be determined whether re-administration of the variant-based or broadly neutralizing epitope-based vaccines can achieve effective cross-variant immunity. Overall, our study suggests that the timely update of the vaccines rather than the durability of the SARS-CoV-2 humoral immunity should be more of our concern.

## Method

### Sample collection

Convalescent sera were collected from a cohort of individuals with prior SARS-CoV-2 infection, collected between January 19, 2021 and January 26, 2021, 305 to 388 days after disease onset. And the illness severity, gender, and age range of donors were recorded. Sera from 10 healthy donors were also obtained. All blood donors provided written informed consent for participation in the study. These human sera protocols were approved by the Ethics Committee of Hubei Provincial Center for Disease Control and Prevention, Wuhan, China (No. 2021-012-01). Sera of the RBD-hFc vaccinated mice (three times immunization) were collected seven weeks post the last immunization from the suborbital venous plexus. The mice sera protocols were approved by the Institutional Animal Care and Use Committee (IACUC) of Center for Animal Experiment, Wuhan University, Wuhan, China (No. WP20210007).

### ELISA detection of SARS-CoV-2 S protein antibodies in the sera

The rapid IgG / IgM test was applied by 2019-nCoV IgG / IgM Detection Kit (Colloidal Gold-Based, Vazyme Biotech Co.,Ltd), which is based on the reactivity of IgG / IgM against SARS-CoV-2 S protein. The anti-RBD IgG level in sera was tested by SARS-CoV-2 (2019-nCoV) Spike RBD Antibody Titer Assay Kit (Sino Biological Inc), which is based on the reactivity of IgG against SARS-CoV-2 RBD. The competitive ability of ACE2 to RBD-binding of sera was tested through SARS-CoV-2 Neutralizing Antibodies Test Kit (ELISA, Wuxi BioHermes Bio & Medical Technology Co., Ltd), which is based on the competition between the neutralizing antibodies in sera and ACE2 to horseradish peroxidase labeled RBD protein (HRP-RBD). These ELISA detections were applied following the manufacturer’s instructions.

### Cell culture

HEK 293T (ATCC, CRL-1168), BHK-21(ATCC, CCL-10) and VERO E6 cells (ATCC, CRL-1586) were cultured in Dulbecco’s medium (DMEM; Gibco) supplemented with 10 % fetal bovine serum (FBS), 2.0mM L-Glutamine, 110 mg/L sodium pyruvate, and 4.5 g/L D-glucose. BHK-21-ACE2 (clone 7) which stably expression human ACE2 was generated based on BHK-21 through lentiviral transduction and maintained with puromycin at 1 μg/ml. I1-Hybridoma (CRL-2700) secreting a monoclonal antibody targeting VSV glycoprotein was cultured in Minimum Essential Medium with Earle’s salts and 2.0 mM L-Glutamine (MEM; Gibco) supplemented with 10 % FBS. All cells were cultured at 37°C in 5 % CO_2_ with regular passage of every 2-3 days.

### Plasmids and protein

The DNA sequences of human codon-optimized S proteins from SARS-COV-2 variants (B.1.1.7, GISAID: EPI-ISL-601443; B.1.351, GISAID: EPI_ISL_678597; P.1, GISAID: EPI_ISL_906075; B.1.525, GISAID: EPI_ISL_1093472) and S protein mutations were commercially synthesized or generated by overlapping-PCR based mutagenesis using pCAGGS-SARS-CoV-2-S-C9 (gifted from Dr. Wenhui Li, National Institute of Biological Science, Beijing,China) as template and cloned into pCAGGS vector with C-terminal 18 aa truncation to improve VSV pseudotyping efficiency(*21*). The plasmid pCAGGS-SARS-CoV-2-RBD-hFc was constructed by inserting the RBD sequence (aa331-525) in to the pCAGGS vector for the expression of RBD-hFc recombinant protein in HEK 293T cells. The proteins were purified by protein A resin (GenScript) following the manufacturer’s instructions.

### SARS-CoV-2 pseudovirus production and titration

The pseudovirus packaged with spike proteins from the WT SARS-CoV-2 and the SARS-CoV-2 variants were produced according to a published protocol with minor modifications(*22*). Briefly, Vero-E6 cells were transfected with plasmids expressing different S proteins through the Lipofectamine 2000 (Biosharp, China). After 24 hrs, the transfected cells were inoculated with VSV-dG-fLuc (1×10^6^ TCID_50_/ml) diluted in DMEM for 5 hrs at 37°C and then replenished with growth medium (DMEM with 10 % FBS) containing anti-VSV-G monoclonal antibody (I1-hybridoma, cultured supernatant, 1:20). 24 hrs later, the SARS-CoV-2 pseudovirus-containing supernatant was harvested and clarified at 3000 r.p.m for 10 min, aliquoted, and frozen at -80°C for storage. The 50 % tissue culture infectious dose (TCID_50_) of pseudovirus was determined by a serial-dilution based infection assay on BHK-21-hACE2 cells and calculated according to the Reed-Muench method(*22, 23*).

### SARS-CoV-2 pseudovirus neutralization assay

The SARS-CoV-2 pseudoviruses were incubated with serial-diluted sera at room temperature for 30 mins in 96-well white flat-bottom culture plates and then mixed with trypsinized BHK-21-hACE2 cells with the density of 2×10^4^/well. After 16 hrs, the medium of the infected cells was removed, and the cells were lysed with 1× Bright-Glo Luciferase Assay reagent (Promega) for chemiluminescence detection with a SpectraMax iD3 Multi-well Luminometer (Molecular devices). The 50 % neutralization dilution titer (NT50) was calculated by GraphPad Prism 7 software with nonlinear regression curve fitting (normalized response, variable slope).

### Authentic SARS-CoV-2 neutralization assay

The SARS-CoV-2 WT strain (IVCAS 6.7512) and B.1.351 strain (NPRC 2.062100001)(*24*) were provided by the National Virus Resource, Wuhan Institute of Virology, Chinese Academy of Sciences, the SARS-CoV-2 B.1.1.7 strain (240108) was provided by HUBEI PROVINCIAL CENTR FOR DISEASE CONTROL AND PREVENTION. All SARS-CoV-2 authentic virus related experiments (S01321010A) were approved by the Biosafety Committee Level 3 (ABSL-3) of Wuhan University, Wuhan Institute of Virology and HUBEI PROVINCIAL CENTR FOR DISEASE CONTROL AND PREVENTION. In brief, sera were serially diluted in culture medium and mixed with 200 TCID_50_ SARS-CoV-2 for 30 mins at room temperature. The mixture was then added to Vero E6 cells in 96-well plates and incubated for 24 hrs, and the cells were fixed with 4 % paraformaldehyde in PBS at room temperature for 1 h, permeabilized with 0.2 % Triton X-100 for 10 mins, and then blocked with 1 % BSA/PBS at 37 °C for 1 h. Cells were subsequently incubated with a mouse monoclonal antibody targeting SARS-CoV/SARS-CoV-2 Nucleocapsid (40143-MM05, Sino Biological) at 1:500 dilution at 37 °C for 1 h, and then incubated with 2 μg/ml of Alexa Fluor 594-conjugated goat anti-mouse IgG antibody (A-11032, Thermo Fisher Scientific) at 37 °C for 1 h. The nucleus was stained with Hoechst 33342. Images were acquired with an inverted fluorescence microscope (DMi8, Leica).

### Statistical analysis

Data are presented as mean values with s.e.m. or s.d.. All statistical analyses were done using GraphPad Prism 7. Differences between two independent samples were evaluated by two-tailed Mann–Whitney U-tests, and *P* < 0.05 is considered statistically significant. Differences between two related samples were evaluated by paired two-tailed t-tests.

## Data Availability

All data are available in the manuscript or extended data.

## Data availability

All data are available in the manuscript or extended data.

## Acknowledgements

This study was supported by grants from the National Science and Technology Major Project (2018YFA0900801), China NSFC projects (32041007 and 32070160), Fundamental Research Funds for the Central Universities (2042021kf0220 and 2042020kf0024), the Advanced Customer Cultivation Project of Wuhan National Biosafety Laboratory (2021ACCP-MS10) and Special Fund for COVID-19 Research of Wuhan University. We are grateful to Beijing Taikang Yicai Foundation for their great support to this work.

## Author contributions

K.L., Y.C., H.Y. and K.C. conceptualized the study design; F.H.M., F.X.Z., X.Y.C., B.H., J.Q.X and Y.Z.J. collected samples; Q.Y.L., Q.X., F.H.M., C.B.M., Z.Z., X.Y.C.,M.G., X.W., Y.H.F., S.S. and C.W.K. did the laboratory tests; Q.Y.L., Q.X., F.H.M., Y.L.L., F.L., L.Z., K.X., C.W.K., F.D., H.Y. and Y.C. analysed the data; Q.Y.L., Q.X., F.H.M., H.Y., Y.C. and K.L. interpreted the results; Q.Y.L., Q.X., F.H.M., H.Y. and Y.C. wrote the initial drafts of the manuscript; Q.Y.L., K.C., H.Y., Y.C. and K.L. revised the manuscript; Y.L.L., F.L., L.Z., K.X., C.W.K., J.Q.X., Y.Z.J. and F.D. commented on the manuscript. All authors read and approved the final manuscript.

## Additional information

Supplementary information which includes Extended data figures and tables are available for this paper.

## Figure legend

**Extended data Fig. 1: The neutralizing activity of the convalescents’ sera to WT-D614G and variants for NT**_**50**_ **determination**.

Convalescents’ sera were serially diluted and then were inoculated with 3×10^5^ TCID50 pseudoviruses as indicated for 30 min at room temperature. BHK-21-hACE2 cells were infected by this mixture, followed by a chemiluminescence detection after 16 hrs. Mean ± s.d. (n=2) are shown. The NT_50_ was calculated by GraphPad Prism 7 software with nonlinear regression curve fitting (normalized response, variable slope). The horizontal dotted lines on each graph indicate 50 % and 0 % neutralization.

**Extended data Table 1: The clinical records and sera reactivity to SARS-CoV-2 of the convalescents in this study**.

