## Supplementary material for "Antibody neutralization to SARS-CoV-2 and variants after one year in Wuhan": Supplemetntal Data

**Extended data Fig. 1: The neutralizing activity of the convalescents' sera to WT-D614G and variants for $NT_{50}$ determination.**

**Extended data Table 1: The clinical records and sera reactivity to SARS-CoV-2 of the convalescents in this study.**

**Extended data Fig. 1**

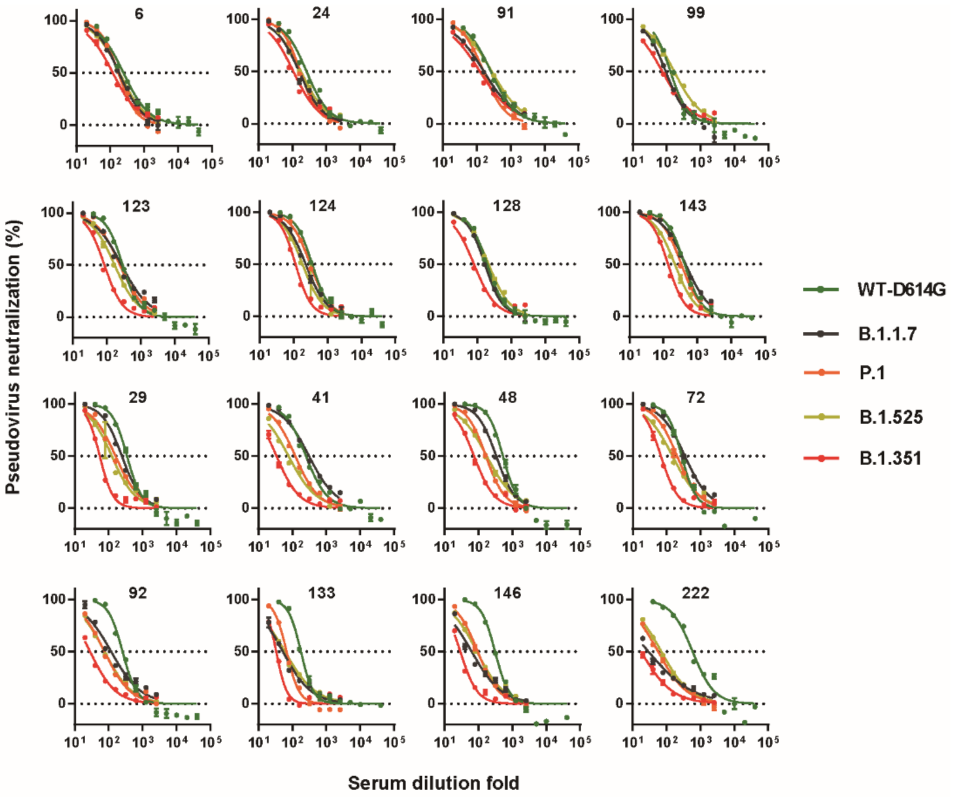


| | | $NT_{50}$ | | | | |
|---|---|---|---|---|---|---|
| | sample number | WT-D614G | England | South Africa | Brazil | Nigeria |
| Group 1 | 6 | 245.5 | 181.6 | 119.3 | 173.8 | 188.5 |
| | 24 | 263 | 158.1 | 107.5 | 171.4 | 180.1 |
| | 91 | 262.4 | 163.4 | 124.5 | 158.4 | 277.7 |
| | 99 | 129 | 93.78 | 74.42 | 93.78 | 166.3 |
| | 123 | 294 | 273 | 85.22 | 243.9 | 170.4 |
| | 124 | 354.1 | 227.2 | 113.9 | 309.4 | 183.2 |
| | 128 | 194.4 | 166.5 | 79.45 | 166.5 | 214.7 |
| | 143 | 421.9 | 453.7 | 137.5 | 338.3 | 217.6 |
| Group 2 | 29 | 262.4 | 163.4 | 124.5 | 158.4 | 277.7 |
| | 41 | 194.4 | 166.5 | 79.45 | 166.5 | 214.7 |
| | 48 | 354.1 | 227.2 | 113.9 | 309.4 | 183.2 |
| | 72 | 245.5 | 181.6 | 119.3 | 173.8 | 188.5 |
| | 92 | 129 | 93.78 | 74.42 | 93.78 | 166.3 |
| | 133 | 294 | 273 | 85.22 | 243.9 | 170.4 |
| | 146 | 421.9 | 453.7 | 137.5 | 338.3 | 217.6 |
| | 222 | 263 | 158.1 | 107.5 | 171.4 | 180.1 |

### Extended data Table 1:

| Sample number | Gender | Age range | Illness severity | Anti-SARS-CoV-2-S IgM | Anti-SARS-CoV-2-S IgG | Anti-RBD IgG level (OD450) | Competitive ability of ACE2 to RBD bingding (%) |
|---|---|---|---|---|---|---|---|
| 1 | male | 35~39 | critical | - | + | 0.954 | 63.6% |
| 2 | female | 40~44 | mild | - | + | 0.498 | 41.7% |
| 3 | male | 55~59 | mild | - | - | 0.608 | 36.2% |
| 4 | female | above 74 | mild | - | + | 0.773 | 82.7% |
| 5 | female | 70~74 | asymptomatic | - | + | 0.606 | 70.1% |
| 6 | female | 65~69 | mild | - | + | 0.83 | 78.5% |
| 7 | male | 50~54 | mild | - | + | 0.608 | 32.0% |
| 8 | female | 65~69 | mild | - | + | 0.948 | 87.9% |
| 9 | female | 50~54 | mild | - | + | 0.58 | 40.4% |
| 10 | male | above 74 | normal | - | + | 0.887 | 71.5% |
| 12 | male | 35~39 | mild | - | - | 0.549 | 45.8% |
| 13 | female | above 74 | mild | - | - | 0.273 | 5.5% |
| 14 | female | 50~54 | normal | - | + | 0.982 | 66.6% |
| 15 | male | 65~69 | mild | - | + | 0.849 | 69.4% |
| 16 | male | 55~59 | normal | - | + | 0.75 | 49.6% |
| 17 | female | 70~74 | mild | - | + | 0.9 | 78.1% |
| 18 | female | 55~59 | severe | - | + | 0.794 | 80.2% |
| 19 | male | 60~64 | mild | - | + | 0.374 | 32.3% |
| 20 | female | 65~69 | severe | - | + | 1.224 | 92.7% |
| 21 | female | 35~39 | mild | - | + | 0.692 | 64.6% |
| 22 | female | 60~64 | mild | - | + | 0.8 | 86.6% |
| 23 | male | 65~69 | severe | - | + | 1.017 | 84.4% |
| 24 | male | above 74 | mild | - | + | 1.118 | 86.7% |
| 25 | female | 60~64 | mild | - | + | 1.145 | 94.7% |
| 26 | male | 35~39 | mild | - | + | 0.551 | 40.0% |
| 27 | female | 70~74 | mild | - | + | 1.188 | 93.3% |
| 28 | female | 60~64 | normal | - | + | 1.191 | 95.8% |
| 29 | female | 60~64 | mild | - | + | 1.367 | 97.1% |
| 30 | female | 60~64 | asymptomatic | - | + | 1.117 | 87.2% |
| 31 | male | 65~69 | severe | - | + | 0.784 | 70.5% |
| 32 | male | 60~64 | severe | - | + | 0.664 | 63.5% |
| 33 | female | 65~69 | mild | - | + | 0.763 | 76.8% |
| 34 | male | 65~69 | normal | - | + | 0.927 | 80.3% |
| 35 | female | 60~64 | normal | - | + | 1.425 | 96.7% |
| 36 | male | 65~69 | severe | - | + | 0.971 | 73.6% |
| 37 | female | 70~74 | mild | - | + | 1.375 | 96.8% |
| 38 | male | 40~44 | normal | - | - | 0.458 | 48.7% |
| 39 | female | 55~59 | normal | - | + | 0.954 | 86.5% |
| 40 | male | 35~39 | normal | - | - | 0.179 | 15.5% |
| 41 | female | 70~74 | normal | - | + | 1.166 | 77.4% |
| 42 | female | 60~64 | severe | - | + | 0.697 | 50.2% |
| 43 | female | 50~54 | mild | - | + | 0.754 | 60.8% |
| 44 | male | 65~69 | severe | - | + | 1.161 | 86.3% |
| 45 | male | above 74 | severe | - | + | 0.432 | 38.8% |
| 46 | female | 55~59 | normal | - | + | 1.151 | 88.5% |
| 47 | male | 55~59 | normal | - | + | 0.901 | 77.6% |
| 48 | female | 55~59 | mild | - | + | 1.293 | 96.7% |
| 49 | female | 50~54 | normal | - | + | 1.327 | 93.4% |
| 50 | male | 70~74 | normal | - | - | 0.275 | 21.3% |
| 51 | female | 45~49 | severe | - | + | 0.782 | 60.8% |
| 52 | male | 55~59 | severe | - | + | 1.17 | 92.1% |
| 53 | male | 70~74 | severe | - | + | 0.402 | 31.4% |
| 54 | female | 65~69 | mild | - | + | 0.458 | 29.1% |
| 55 | female | 65~69 | severe | - | + | 0.632 | 62.6% |
| 56 | female | 40~44 | normal | - | + | 1.378 | 97.1% |
| 57 | male | 45~49 | severe | - | - | 0.339 | 32.4% |
| 58 | male | 55~59 | mild | - | + | 1.081 | 92.7% |
| 59 | male | 20~29 | asymptomatic | - | - | 0.125 | 17.0% |
| 60 | male | 65~69 | normal | - | + | 1.328 | 96.3% |
| 61 | female | 60~64 | critical | - | - | 0.689 | 64.6% |
| 62 | male | 55~59 | severe | - | + | 1.092 | 77.7% |
| 63 | female | 60~64 | normal | - | + | 0.866 | 81.6% |
| 64 | female | 60~64 | mild | - | - | 0.546 | 38.2% |

| Sample number | Gender | Age range | Illness severity | Anti-SARS-CoV-2-S IgM | Anti-SARS-CoV-2-S IgG | Anti-RBD IgG level (OD450) | Competitive ability of ACE2 to RBD bingding (%) |
|---|---|---|---|---|---|---|---|
| 65 | female | 60~64 | severe | - | + | 0.89 | 71.6% |
| 66 | female | 55~59 | normal | - | + | 0.763 | 65.1% |
| 67 | female | 55~59 | severe | - | + | 0.743 | 64.7% |
| 68 | male | above 74 | mild | - | + | 0.847 | 63.4% |
| 69 | male | 30~34 | normal | - | + | 0.531 | 56.0% |
| 70 | male | 70~74 | normal | - | + | 0.967 | 76.5% |
| 71 | female | 70~74 | normal | - | + | 0.667 | 51.6% |
| 72 | female | 40~44 | normal | - | + | 1.293 | 94.3% |
| 73 | male | 50~54 | mild | - | + | 0.254 | 12.4% |
| 74 | female | 50~54 | normal | - | - | 0.199 | 18.9% |
| 75 | male | 55~59 | severe | - | + | 0.544 | 63.4% |
| 76 | male | 65~69 | normal | - | + | 0.351 | 40.5% |
| 77 | female | 20~29 | mild | - | - | 0.376 | 35.8% |
| 78 | female | 65~69 | mild | - | + | 1.044 | 83.0% |
| 79 | male | 50~54 | mild | - | + | 0.211 | 56.6% |
| 80 | female | 55~59 | normal | - | + | 0.763 | 83.8% |
| 81 | female | 65~69 | mild | - | + | 0.726 | 56.2% |
| 82 | female | 60~64 | mild | - | + | 0.606 | 47.4% |
| 83 | female | 55~59 | normal | - | + | 1.146 | 66.8% |
| 84 | male | 45~49 | mild | - | + | 0.672 | 48.3% |
| 85 | female | 45~49 | normal | - | - | 0.401 | 46.5% |
| 86 | male | 60~64 | severe | - | + | 0.616 | 47.5% |
| 87 | female | 45~49 | mild | + | + | 1.467 | 94.9% |
| 88 | male | 45~49 | normal | - | - | 0.429 | 33.3% |
| 89 | female | 50~54 | mild | - | + | 0.925 | 69.5% |
| 90 | female | 50~54 | normal | - | + | 0.855 | 76.9% |
| 91 | male | 55~59 | mild | + | + | 0.901 | 65.7% |
| 92 | female | 55~59 | critical | - | + | 1.058 | 96.7% |
| 93 | female | 50~54 | mild | - | + | 1.326 | 96.4% |
| 94 | male | 55~59 | normal | - | + | 0.837 | 92.3% |
| 95 | male | 55~59 | severe | - | + | 1.272 | 95.0% |
| 96 | female | 50~54 | normal | - | + | 0.683 | 34.2% |
| 97 | male | 20~29 | mild | - | + | 0.826 | 60.2% |
| 98 | male | 55~59 | severe | - | + | 1.008 | 90.3% |
| 99 | female | 60~64 | mild | - | + | 0.378 | 53.2% |
| 100 | female | 50~54 | normal | - | + | 0.223 | 17.0% |
| 101 | male | 40~44 | normal | - | - | 0.638 | 24.6% |
| 102 | female | 45~49 | normal | - | - | 0.546 | 43.0% |
| 103 | female | 55~59 | asymptomatic | - | - | 0.376 | 4.2% |
| 104 | male | 50~54 | severe | - | + | 0.908 | 82.1% |
| 105 | female | 50~54 | normal | - | + | 0.686 | 59.4% |
| 106 | male | 40~44 | critical | + | + | 0.46 | 30.6% |
| 107 | female | 40~44 | normal | - | + | 0.777 | 54.6% |
| 108 | male | 65~69 | mild | - | + | 1.136 | 88.5% |
| 109 | male | 20~29 | mild | - | + | 0.983 | 80.5% |
| 110 | male | 60~64 | mild | - | + | 0.731 | 19.6% |
| 111 | female | 65~69 | normal | - | + | 0.721 | 35.6% |
| 112 | female | 35~39 | severe | - | + | 0.516 | 21.7% |
| 113 | male | 55~59 | mild | - | + | 1.041 | 83.3% |
| 115 | female | 20~29 | mild | - | - | 0.58 | 61.4% |
| 116 | male | 55~59 | severe | - | - | 0.49 | 38.0% |
| 117 | male | 55~59 | mild | - | - | 0.624 | 39.8% |
| 118 | female | 65~69 | mild | - | + | 0.051 | 62.6% |
| 119 | female | 35~39 | mild | - | + | 0.989 | 16.8% |
| 120 | female | 65~69 | severe | - | + | 0.936 | 88.8% |
| 121 | female | 50~54 | normal | - | + | 0.664 | 76.6% |
| 122 | female | 20~29 | mild | - | + | 0.829 | 85.1% |
| 123 | female | 50~54 | normal | - | + | 0.919 | 95.6% |
| 124 | male | 55~59 | mild | - | + | 1.184 | 96.3% |
| 125 | male | 60~64 | normal | - | + | 0.58 | 43.0% |
| 126 | male | 65~69 | normal | - | - | 0.242 | 21.4% |
| 127 | male | 60~64 | normal | - | + | 0.512 | 46.0% |
| 128 | female | 65~69 | normal | - | + | 0.82 | 74.6% |
| 129 | male | 60~64 | mild | - | - | 0.125 | 2.2% |
| 130 | male | 60~64 | severe | - | + | 0.729 | 49.0% |
| 131 | male | 70~74 | mild | - | - | 0.322 | 13.6% |

| Sample number | Gender | Age range | Illness severity | Anti-SARS-CoV-2-S IgM | Anti-SARS-CoV-2-S IgG | Anti-RBD IgG level (OD450) | Competitive ability of ACE2 to RBD bingding (%) |
|---|---|---|---|---|---|---|---|
| 132 | female | 70~74 | mild | - | + | 0.642 | 60.5% |
| 133 | male | 70~74 | mild | - | + | 1.068 | 50.5% |
| 134 | male | 50~54 | normal | - | + | 0.702 | 61.2% |
| 135 | female | 55~59 | mild | - | - | 0.47 | 42.2% |
| 136 | male | 45~49 | mild | - | - | 0.246 | 15.1% |
| 137 | female | 55~59 | normal | - | - | 0.894 | 53.6% |
| 138 | female | 65~69 | mild | - | + | 0.935 | 60.5% |
| 139 | male | 30~34 | mild | - | + | 0.318 | 21.4% |
| 140 | male | 60~64 | normal | - | + | 1.024 | 72.8% |
| 141 | male | 35~39 | normal | - | - | 0.549 | 46.4% |
| 142 | female | 40~44 | normal | - | + | 1.18 | 94.4% |
| 143 | female | 65~69 | mild | - | + | 1.191 | 95.5% |
| 144 | female | 60~64 | normal | - | + | 0.376 | 15.1% |
| 145 | male | 60~64 | mild | - | - | 0.727 | 62.6% |
| 146 | female | 55~59 | mild | - | + | 1.177 | 96.8% |
| 147 | female | 65~69 | severe | - | + | 0.788 | 79.7% |
| 148 | female | 55~59 | mild | - | + | 1.15 | 96.1% |
| 149 | female | 50~54 | normal | - | - | 0.219 | 13.0% |
| 150 | female | 30~34 | mild | - | + | 0.553 | 47.4% |
| 151 | male | 40~44 | severe | - | + | 0.64 | 79.0% |
| 152 | male | 60~64 | mild | - | + | 0.8 | 70.5% |
| 153 | male | 60~64 | asymptomatic | - | + | 0.265 | 22.0% |
| 154 | male | 30~34 | normal | - | + | 0.598 | 49.4% |
| 155 | female | 40~44 | normal | - | + | 1.12 | 94.0% |
| 156 | female | 55~59 | normal | - | - | 0.234 | 40.1% |
| 157 | female | 20~29 | mild | - | + | 1.186 | 96.3% |
| 158 | male | 30~34 | mild | - | - | 0.806 | 55.4% |
| 159 | female | 55~59 | mild | - | + | 0.614 | 45.3% |
| 160 | female | 55~59 | normal | - | + | 0.956 | 79.6% |
| 161 | male | above 74 | mild | - | - | 0.591 | 61.6% |
| 162 | female | above 74 | normal | - | + | 0.516 | 46.0% |
| 163 | female | 70~74 | mild | + | + | 0.411 | 45.7% |
| 164 | female | 65~69 | normal | - | + | 0.805 | 77.9% |
| 165 | male | 70~74 | severe | - | + | 0.476 | 31.6% |
| 166 | female | 70~74 | critical | - | + | 0.86 | 74.5% |
| 167 | female | 60~64 | severe | - | + | 0.585 | 52.2% |
| 168 | female | 45~49 | critical | - | + | 0.277 | 12.0% |
| 169 | female | 40~44 | mild | - | + | 1.053 | 80.1% |
| 170 | male | 65~69 | severe | - | + | 0.478 | 31.1% |
| 171 | female | 65~69 | severe | - | + | 0.361 | 22.8% |
| 172 | male | 40~44 | severe | - | + | 0.688 | 34.7% |
| 173 | male | above 74 | normal | - | + | 0.598 | 33.2% |
| 174 | male | 65~69 | mild | - | + | 0.316 | 17.8% |
| 175 | male | 65~69 | mild | - | + | 0.751 | 77.4% |
| 176 | female | 65~69 | mild | - | + | 0.706 | 55.6% |
| 177 | female | 70~74 | mild | - | + | 1.218 | 79.5% |
| 178 | female | 70~74 | mild | - | + | 1.064 | 54.0% |
| 179 | male | 30~34 | mild | - | + | 0.68 | 29.9% |
| 180 | female | 55~59 | normal | - | + | 1.335 | 78.7% |
| 181 | male | 70~74 | severe | - | - | 0.259 | 0.0% |
| 182 | female | 60~64 | normal | - | + | 1.451 | 62.8% |
| 183 | female | above 74 | normal | - | + | 0.799 | 23.5% |
| 184 | male | 50~54 | mild | - | + | 0.604 | 10.7% |
| 185 | male | 65~69 | normal | - | + | 1.641 | 80.7% |
| 186 | male | 55~59 | mild | - | + | 0.638 | 27.2% |
| 187 | female | 65~69 | normal | - | - | 0.481 | 16.0% |
| 188 | male | 35~39 | mild | - | + | 0.607 | 40.7% |
| 189 | male | 60~64 | severe | - | + | 1.106 | 67.7% |
| 190 | female | 60~64 | normal | - | + | 0.828 | 58.2% |
| 191 | male | 60~64 | critical | - | - | 0.79 | 33.4% |
| 192 | male | 50~54 | normal | - | + | 0.775 | 45.0% |
| 193 | female | 50~54 | mild | - | + | 0.65 | 9.7% |
| 194 | male | 30~34 | normal | - | + | 0.801 | 40.3% |
| 195 | male | 35~39 | mild | - | - | 0.331 | 0.0% |
| 196 | male | 70~74 | mild | - | + | 1.267 | 63.9% |
| 197 | female | 65~69 | severe | - | - | 0.2 | 2.8% |

| Sample number | Gender | Age range | Illness severity | Anti-SARS-CoV-2-S IgM | Anti-SARS-CoV-2-S IgG | Anti-RBD IgG level (OD450) | Competitive ability of ACE2 to RBD bingding (%) |
|---|---|---|---|---|---|---|---|
| 198 | female | 55~59 | normal | - | + | 1.692 | 88.9% |
| 199 | female | 20~29 | normal | - | + | 0.783 | 23.0% |
| 200 | male | 60~64 | severe | - | + | 1.204 | 73.8% |
| 201 | male | 60~64 | severe | - | + | 0.942 | 55.6% |
| 202 | female | 60~64 | mild | - | + | 0.879 | 47.9% |
| 203 | female | 60~64 | critical | - | + | 1.256 | 87.1% |
| 204 | male | 30~34 | severe | - | + | 1.028 | 70.9% |
| 205 | female | 60~64 | critical | - | + | 0.716 | 45.7% |
| 206 | male | 65~69 | normal | - | + | 0.826 | 65.3% |
| 207 | male | 65~69 | severe | - | + | 0.969 | 83.5% |
| 208 | female | above 74 | mild | - | - | 0.354 | 30.4% |
| 209 | female | 65~69 | mild | - | - | 0.499 | 16.3% |
| 210 | female | 60~64 | severe | - | + | 0.932 | 59.5% |
| 211 | female | 55~59 | severe | - | - | 0.717 | 41.1% |
| 212 | female | 35~39 | normal | - | + | 0.772 | 54.0% |
| 213 | male | 55~59 | asymptomatic | - | + | 1.362 | 92.1% |
| 214 | female | 60~64 | normal | - | + | 1.503 | 84.7% |
| 215 | female | 65~69 | mild | - | + | 0.964 | 43.3% |
| 216 | male | 65~69 | critical | - | - | 0.825 | 21.7% |
| 217 | male | 65~69 | severe | - | + | 1.321 | 67.6% |
| 218 | female | 65~69 | normal | - | + | 1.226 | 83.0% |
| 219 | male | 65~69 | normal | - | + | 1.181 | 78.7% |
| 220 | male | 60~64 | severe | - | - | 0.471 | 31.5% |
| 221 | female | 30~34 | mild | - | - | 0.845 | 53.5% |
| 222 | male | 65~69 | normal | - | + | 1.597 | 92.2% |
| 223 | male | 65~69 | normal | - | + | 1.387 | 86.9% |
| 224 | male | 70~74 | severe | - | + | 0.97 | 76.3% |
| 225 | female | 70~74 | normal | - | + | 0.668 | 47.6% |
| 226 | female | 65~69 | mild | - | + | 0.553 | 52.5% |
| 227 | male | 70~74 | mild | - | - | 0.975 | 63.5% |
| 228 | female | above 74 | asymptomatic | - | + | 0.739 | 68.3% |
| 229 | male | 65~69 | severe | - | + | 0.871 | 87.8% |
| 230 | male | 55~59 | asymptomatic | - | + | 0.752 | 40.2% |
| 231 | male | 30~34 | normal | - | - | 0.952 | 53.6% |
| 232 | male | above 74 | mild | - | + | 0.451 | 1.9% |
| 233 | female | 55~59 | mild | - | + | 0.733 | 45.6% |
| 234 | male | 65~69 | mild | - | - | 0.558 | 21.1% |
| 235 | female | 40~44 | mild | - | - | 0.237 | 0.0% |
| 236 | male | 20~29 | normal | - | - | 0.185 | 0.9% |
| 237 | male | 50~54 | normal | - | + | 0.775 | 72.8% |
| 238 | male | 45~49 | mild | - | + | 1.152 | 84.6% |
| 239 | male | 70~74 | mild | + | + | 1.245 | 82.6% |
| 240 | female | 70~74 | mild | - | + | 0.695 | 52.9% |
| 241 | male | 60~64 | normal | - | - | 0.23 | 12.8% |
| 242 | female | 20~29 | mild | - | + | 1.354 | 95.6% |
| 243 | female | 60~64 | mild | - | - | 0.218 | 10.2% |
| 244 | male | 70~74 | mild | - | + | 0.914 | 63.0% |
| 245 | male | 45~49 | mild | - | + | 0.524 | 25.8% |
| 246 | male | 60~64 | mild | - | + | 0.735 | 45.7% |
| 247 | female | 60~64 | mild | - | + | 0.866 | 48.6% |
| 248 | female | above 74 | normal | - | - | 0.119 | 0.0% |
| 249 | male | 60~64 | normal | - | + | 0.792 | 71.5% |
| 250 | female | 70~74 | mild | - | + | 1.081 | 86.3% |
| NC-1 | male | 30~34 | uninfected | - | - | 0.119 | untested |
| NC-2 | male | 30~34 | uninfected | - | - | 0.178 | untested |
| NC-3 | male | 20~29 | uninfected | - | - | 0.124 | untested |
| NC-4 | female | 20~29 | uninfected | - | - | 0.201 | untested |
| NC-5 | female | 20~29 | uninfected | - | - | 0.048 | untested |
| NC-6 | male | 35~39 | uninfected | - | - | 0.175 | untested |
| NC-7 | female | 40~44 | uninfected | - | - | 0.101 | untested |
| NC-8 | female | 55~59 | uninfected | - | - | 0.243 | untested |
| NC-9 | female | 60~64 | uninfected | - | - | 0.153 | untested |
| NC-10 | male | 45~49 | uninfected | - | - | 0.13 | untested |
